# Highly Pathogenic Avian Influenza Vaccines: challenges for innovation, technological development and pandemic preparedness

**DOI:** 10.1101/2023.05.31.23290790

**Authors:** Cristina Possas, Ernesto TA Marques, Alessandra Oliveira, Suzanne Schumacher, Flavia Mendes, Marilda M Siqueira, Adelaide Antunes, Akira Homma

## Abstract

**Introduction:** Highly Pathogenic Avian Influenza (HPAI) pandemic potential is a critical issue. Outbreaks of H5N1 Avian Flu, causing mass die-offs in poultry and other birds, are now affecting mammals from otters to dolphins worldwide. H5N1 and other HPAI viruses have so far caused rare human infections. However, if they evolve to spread between people, they could trigger major outbreaks. The main question is: how to overcome bottlenecks in HPAI vaccine development to accelerate scale-up and implement an effective global Vaccine Preparedness System?

**Methods:** From a patent landscape approach, we identified breakthroughs and gaps in HPAI vaccine development for the search period (2010-2021), discussing technological strategies in patent filings, including universal vaccine patent documents.

**Results:** Very few patent documents for human HPAI vaccines were retrieved: from 49 deposits in the search period, only 18 were in active status (37%), with few breakthroughs and in early stages of development. These results indicate a technological lag in HPAI vaccine development and highlight the constraints for scaling-up these vaccines when they will be needed. Current egg-based processes are laborious and too difficult to scale-up in the case of a pandemic. Concerning mRNA technology, there are still issues on the duration and level of protection mRNA vaccines might induce. Vaccinating farm animals is not sufficient to protect the population: if not properly executed, the virus could continue to circulate at a low level, increasing the chance of mutations and adaptation to humans. Universal Influenza vaccines would be key, but we identified only few (20 in the search period) patents for these vaccines, still in early stages of development. In addition, it is the impact immunological memory would have on the development of protective immunity to a broad-spectrum of viruses, due to original antigenic sin mechanism.

**Discussion:** This study identified critical barriers to overcome in HPAI vaccine development, improving efficacy and reducing costs. It highlights the urgent need for an effective global Vaccine Preparedness model, supported by Genomic, Antigenic and Epidemiological Surveillance, an Innovation Fund and Public-Private Partnerships. We strongly support WHO in coordinating a Global Initiative for Influenza Pandemic Preparedness.

## 1 Introduction

Influenza viruses are complex and constantly changing, instigating a broad range of scientific issues due to their highly changeable nature and dynamic profile. The four types of influenza virus are A, B, C and D. Different from endemic Seasonal A Influenza, Highly Pathogenic A Influenza (HPAI) can spread easily among animals and potentially cause a future pandemic, due to the virus’ capacity to infect people and to sustain human-to-human transmission (1, 2).

This Influenza A virus is considered one of the most important to public health due to its very high pandemic potential. Influenza A is often classified based on host′s species (Figure 1), such as swine influenza and avian influenza. Influenza B virus can also cause seasonal epidemics and circulates among humans and can infect seals. Influenza C virus is normally mild and can infect humans and pigs. And finally, influenza D virus which it cannot infect human, and primarily infect cattle (1). These Flu viruses change in two different ways (“antigenic drift” – small changes or mutations in viral genes and “antigenic shift” – an abrupt, major viral change, which can happen if an Influenza virus from an animal population evolves to infects humans (“spill-over”), as indicated in Figure 1.

**Figure 1.**
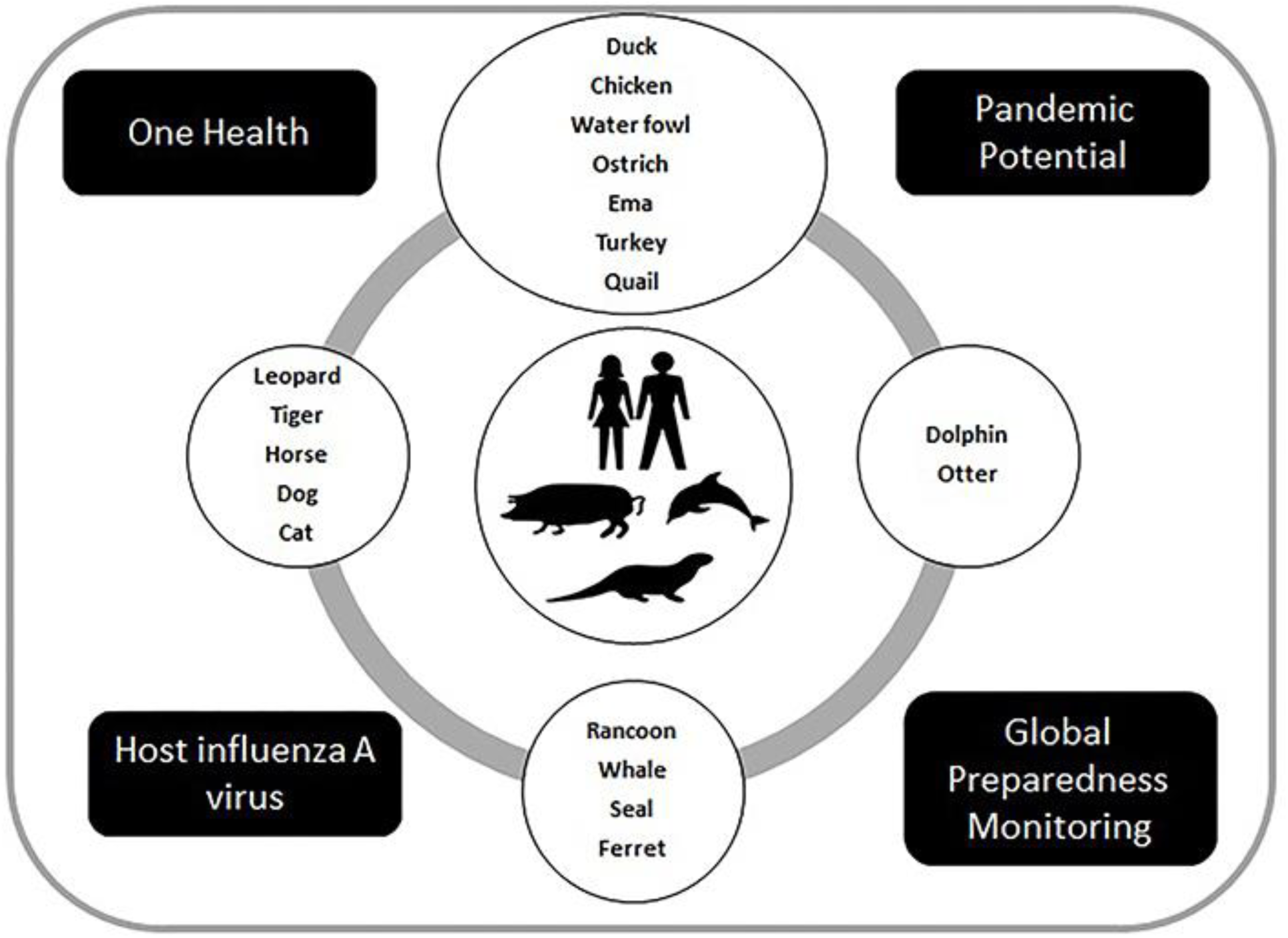
Hosts influenza A virus and the pandemic potential Source: Elaborated by the authors based on data from (2).

Human infections with highly-pathogenic bird flu viruses have occurred in most cases after unprotected contact with infected birds (wild birds/domestic poultry) or contaminated surfaces. Nevertheless, some infections have emerged where direct contact with infected birds or their environments was not identified. Human-to-human transmission has been so far very rare and when it happened, it has only spread to few people. However, because of the possibility that bird Influenza viruses could mutate and spread easily between people, in outbreaks and even in a human pandemic, genetic and immune type monitoring for human infection and person-to-person spread is extremely important for public health (3, 4). Concerning potentially pandemic strains, Asian lineage H7N9 and highly-pathogenic avian influenza Asian lineage H5N1 viruses have been responsible for most human illness from bird Influenza viruses worldwide to date, including the most serious illnesses and illness with the highest mortality. With regard to the H5N1 strain producing human disease, to date more than 50% of the diagnosed infections have been fatal (3). Although it should be recognized that there is a variety of genotypes and immunotypes of H7N9 and H5N1 strains with relatively low severity and lethality, the highly pathogenic and pandemic potential of these two lineages has been a matter of global concern by public health authorities.

An Influenza pandemic is thus a global outbreak of a new Influenza A virus and happens when novel Influenza A viruses emerge and gain the ability to infect humans easily and spread from person to person in an efficient and sustained way (5). Although Influenza usually emerges as seasonal outbreaks, with pandemics being relatively rare, with five Influenza pandemics since the beginning of last century, the possibility of emergence of a new Influenza A pandemic has been a matter of great global concern due to their potential for enormous social and economic impacts. This concern has increased in the current global scenario of environmental changes, mass production of animal food and high-density populated cities that facilitate the emergence and transmission of severe respiratory syndromes, as demonstrated by the COVID-19 pandemic.

*“An influenza pandemic is a global epidemic caused by a new influenza virus to which there is a little or no pre-existing immunity in the human population”* (6). Highly-pathogenic influenza viruses are associated with rapid global spread/severe disease and they have been responsible for many deaths. Five Influenza pandemics occurred since 1918: H1N1 (1918–1920) when one third of the world population became infected and at least 50 million people died), H2N2 (1957–1958), H3N2 (1968–1969), a much milder H1N1 (1977–1978) and a recent H1N1 pandemic (2009) which caused between 100, 000 and 400,000 deaths globally in the first year (6, 7). Moreover, H5N1, a highly pathogenic A influenza has emerged in 1997, circulating worldwide in avian populations with reports of human cases and deaths. This strain was observed in 16 countries (649 patients whom almost 60% have died) and it was first identified in 1997 in Hong Kong. In Figure 2 we provide a global overview of confirmed human cases of Avian Influenza H5N1 reported to WHO from 1997 to 2022.

**Figure 2.**
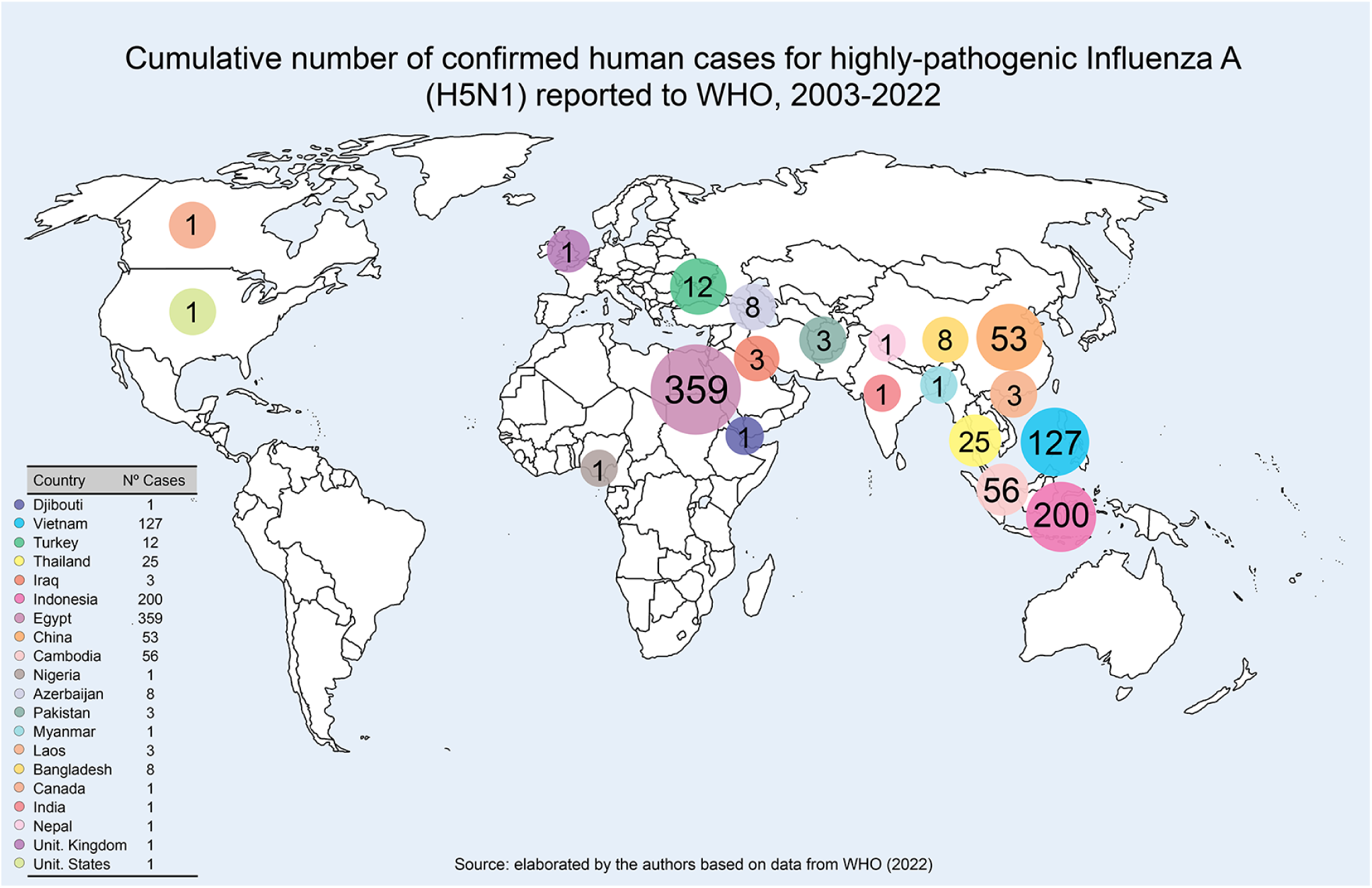
Reported global human cases with highly pathogenic avian influenza A (H5N1 (HPAI H5N1)) by country, 1997-2022 Source: Elaborated by the author based on WHO (2022) (4)

At the dawn of the COVID-19 pandemic, there has been an increased recognition that more attention needs to be paid to global preparedness and human vaccines against highly pathogenic respiratory virus. Flu viruses can mutate and gain the ability to be transmitted from animals to humans: a zoonotic spillover process that can spread among people and cause a pandemic (7, 8). A significant amount of research is now concentrated in this area. The Viral Genomic Project (VGP) is one of the scientific responses to this concerning global scenario, aiming the detection and identification of zoonotic viral threats to promote food security and human health. According to VGP, from 1,67 million unknown viruses from birds and mammals, estimates indicate that 631, 000 to 827, 000 of them can infect humans with a high risk (99%) of viral emergence and spread from ′spill-over (9–12). Considering the severity of this global scenario, it is imperative to better understand how to respond to future outbreaks with pandemic potential, which severely compromise peoples’ well-being and global health security.

From this perspective, academia and governments must urgently collaborate in the search for innovative vaccine development, integrating Vaccine Preparedness into an effective Global Pandemic Preparedness Plan (13). According to Krammer & Palese (2015) (14), the processes of influenza vaccine production have increased significantly since the 2009 H1N1 pandemic. In fact, the authors presented virus vaccines technologies in different development stages, i.e., licensed, clinical and preclinical stages. For the present study, it is important to note the technologies used in broad/mainly universal breadth of protection vaccines: Modified Vaccinia Virus Ankara (MVA)-vectored vaccines, M2e, Chimeric and Headless Haemagglutinin. In this way, novel technologies promote a detailed human immune response analysis on influenza A and B viruses, enabling the development of an immunogenic, safe and universal vaccine. The Universal Vaccine, defined by Dr. Peter Palese^1^ is ‘a vaccine that you don’t have to take every year, maybe only once in a lifetime” (15). However, a more realistic goal is the prevention of severe disease and death. For this, the biological basis must be also explored to vaccines’ development, such as the increase of immunogenicity of avian influenza vaccines. For this goal, adjuvants can be used in the development process (16).

## 2 Materials and methods

For the purpose of this study, the following schematic applied method was used to retrieve the Launched influenza A Vaccines and data about Technological focus on patent landscape and Universal influenza vaccines, as follows (Figure 3).

**Figure 3.**
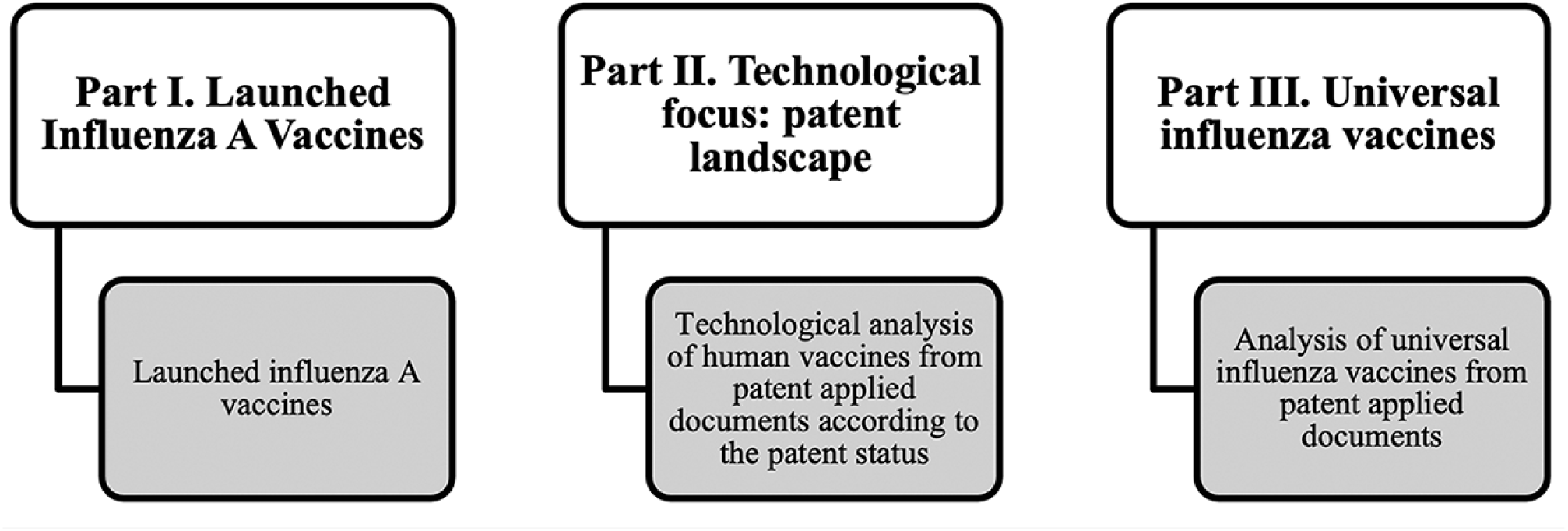
Schematic applied method (2010–2021) Source: Elaborated by the authors

### 2.1 Launched Influenza A Vaccines (Cortellis Drug Discovery Intelligence – CDDI)

Data on launched influenza A vaccines between 2010-2021 were extracted from Cortellis Drug Discovery Intelligence (CDDI) ^2^ (from Clarivate™). The methodology of data collection was based on the developed strategy ^3^ to retrieve the vaccines against influenza A launched 2010-2021 (17) (Figure 3).

### 2.2 Technological analysis of human vaccines from patent applied documents (Derwent Innovation)

According to the methodology, the search was made in the Derwent Innovation database between 2010-2021; The strategy searched for descriptors in the title/abstract/claims in applied patent documents: (Pandemic Potential or H1N1 OR Spanish flu OR swine flu pandemic OR swine flu OR H2N2 OR Asian flu OR H3N2 OR Hong Kong flu OR H5N1) matching the Derwent Manual Code^4^ (B14-S11 – Description: (Vaccine general)) AND (vaccine* or vaccination)). Based on the results, the applied documents from the top applicants were discussed with a focused technological review in a chronological way from the priority year. All the applied documents were retrieved regardless of their patent status, considering the active status only ^5^.

In order to identify the Legal Status of the retrieved applied patent documents two databases were used: Lens and Indian Patent Advanced Search System (inPASS).

Lens is a free database, which contains information on more than 144.3 million patent records, in addition to 252 million academic papers (18). To identify the status of each application, the Publication Number was used.

The identification of the status of Indian documents was carried out using the inPASS database (19) launched in 2015 (21) by the Controller General of Patents Designs and Trademark, subordinated to the government of India. The search was performed using the original title of the patent document and the filing date.

### 2.2 Universal vaccines against influenza

The method has been conceived to retrieve human universal influenza vaccines following a two-step process: First, the search was performed on the Derwent Innovation Index database^6^ according to the strategy developed from Part II (“Technological analysis of human vaccines from patent applied documents”). Second: This simple and practical method allowed us to retrieve the documents related to “Universal Vaccines”, which are effective against all influenza strains. For this purpose, the terms^7^ “universal vaccine”, “universal influenza vaccine”, “universal flu vaccines”, “pan-influenza vaccine” and “pan-flu vaccine” were adopted in different fields of patent applied documents, i.e., title, abstract and claims. Influenza vaccines for veterinary use were excluded from the analysis (Figure 3).

## 3 Results

### 3.1 Launched Influenza Vaccines

The method conceived here was sufficiently to retrieve the launched/registered vaccines in 2010-2021period. Nineteen (19) drugs & biologics (launched influenza A vaccines) ^8^ were recorded. A key finding emerged on the drug type classification: most of them were biotechnologies (16), while a few numbers (3) were classified as peptides. Considering, the country where the vaccine was launched, the United States is the country with the leading number of vaccines in different development statuses (9), all of them launched/registered drugs & biologics, followed by India, Republic of Korea and United Kingdom (each one with 3 launched vaccines). Meanwhile the top organizations were SANOFI (from France, with 4 products), GLAXOSMITHKLINE (from United Kingdom, 3) and SEQIRUS (from United Kingdom, 2) and SK BIOSCIENCE (from South Korea, with also 2 products) (17).

Furthermore, it is important to highlight the low number of launched vaccines against influenza: 19 products in eleven years (2010–2021). In fact, the developed method retrieved a low number of launched^9^ influenza vaccines in the studied period: 2010 (4 launched influenza vaccines); 2013 (5); 2015 (1); 2016 (5); 2017 (3) and 2018 (1). Table 1 provides an overview of the most recent vaccines, i. e., the vaccines launched in 2017 and 2018:

**Table 1.**
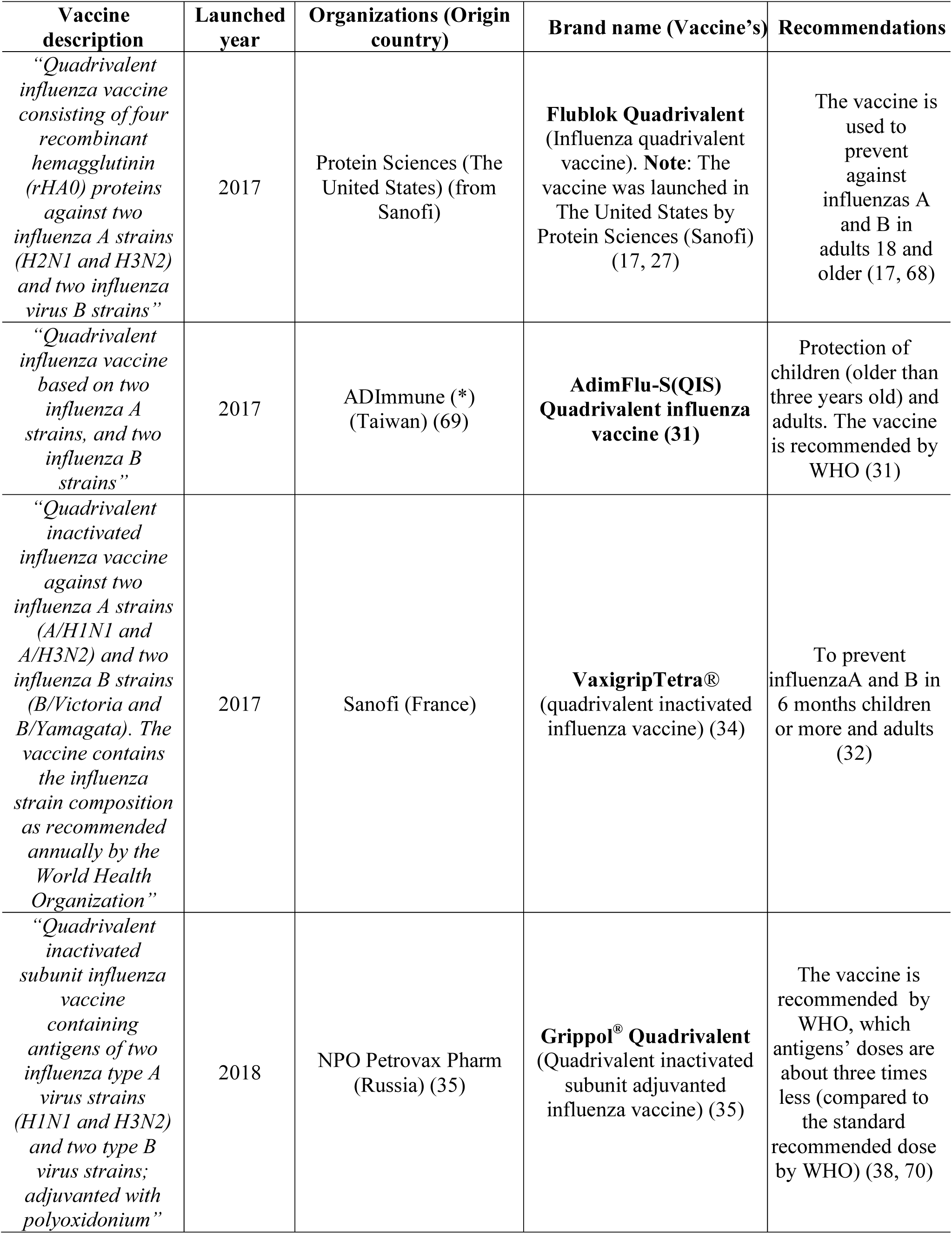
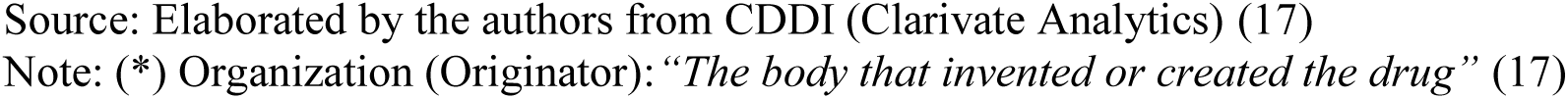
Description of the most recent launched influenza vaccines

| **Vaccine description** | **Launched year** | **Organizations (Origin country)** | **Brand name (Vaccine's)** | **Recommendations** |
| --- | --- | --- | --- | --- |
| *“Quadrivalent influenza vaccine consisting of four recombinant hemagglutinin (rHA0) proteins against two influenza A strains (H2N1 and H3N2) and two influenza virus B strains”* | 2017 | Protein Sciences (The United States) (from Sanofi) | **Flublok Quadrivalent** (Influenza quadrivalent vaccine). **Note:** The vaccine was launched in The United States by Protein Sciences (Sanofi) (17, 27) | The vaccine is used to prevent against influenzas A and B in adults 18 and older (17, 68) |
| *“Quadrivalent influenza vaccine based on two influenza A strains, and two influenza B strains”* | 2017 | ADImmune (*) (Taiwan) (69) | **AdimFlu-S(QIS) Quadrivalent influenza vaccine (31)** | Protection of children (older than three years old) and adults. The vaccine is recommended by WHO (31) |
| *“Quadrivalent inactivated influenza vaccine against two influenza A strains (A/H1N1 and A/H3N2) and two influenza B strains (B/Victoria and B/Yamagata). The vaccine contains the influenza strain composition as recommended annually by the World Health Organization”* | 2017 | Sanofi (France) | **VaxigripTetra®** (quadrivalent inactivated influenza vaccine) (34) | To prevent influenzaA and B in 6 months children or more and adults (32) |
| *“Quadrivalent inactivated subunit influenza vaccine containing antigens of two influenza type A virus strains (H1N1 and H3N2) and two type B virus strains; adjuvanted with polyoxidonium”* | 2018 | NPO Petrovax Pharm (Russia) (35) | **Grippol® Quadrivalent** (Quadrivalent inactivated subunit adjuvanted influenza vaccine) (35) | The vaccine is recommended by WHO, which antigens' doses are about three times less (compared to the standard recommended dose by WHO) (38, 70) |

Source: Elaborated by the authors from CDDI (Clarivate Analytics) (17)
Note: (\*) Organization (Originator): “*The body that invented or created the drug*” (17)

#### 3.1.1 Flublok®Quadrivalent(Launched in 2017: USA)

The influenza vaccine called Flubok® Quadrivalent was developed by PROTEIN SCIENCES CORPORATIONS (US), a SANOFI company (France), which is an important developer of recombinant vaccine (27, 28). The vaccine is approved for persons 18 years or older and it is used to immunize against influenza A and B (29). The quadrivalent vaccine is available for 2021-2022 influenza season and is produced using recombinant technology. In fact, in the manufacturing process, the recombinant flu vaccines do not use chicken eggs or flu virus (egg-free flu vaccine). This has significant benefits in terms of speed of the influenza manufacturing process. Since the product is not dependent on egg supply, it can be produced faster when it is compared to other egg-based vaccines, what is crucial in a pandemic scenario. Another particular advantage arising from how the recombinant technology can affect how the final vaccine works, due to the production process avoids virus mutations that can occur when eggs are needed to viruses growth (30).

#### 3.1.2 AdimFlu-S® (QIS) - quadrivalent influenza vaccine (Lanched in 2017 in Taiwan)

The quadrivalent vaccine, without any live virus particles (H1N1, H3N2, Yamagata and Victoria types B lineages) protects children (older than three years old) and adults for 6 – 12 months. To prepare the vaccine, the influenza virus is cultured in embryonated eggs, and after it is purified, splitted and inactivated (31).

#### 3.1.3 VaxigripTetra®: Quadrivalent inactivated influenza vaccine (Launched in 2017 in Europe)

The vaccine against influenza from Sanofi Pasteur (France) contains four-strain influenza vaccine (influenza A strains:H1N1 AND H3N2 and influenza B strains: Victoria and Yamagata). It is indicated for 6 months old children or more and adults. Available in 20 European markets, it will be planned to be launched in the future all around the world. It (32, 33). In fact, the vaccine is a quadrivalent split-virion vaccine as a well-established record of Vaxigrip® (a trivalent split-virion flu vaccine) (34).

#### 3.1.4 Grippol® Quadrivalent: quadrivalent inactivated subunit adjuvanted influenza vaccine (Lauched in 2018 in Russia)

Grippol® Quadrivalent was developed by Petrovax Pharma, a Russian full-cycle bio-pharmaceutical company. The product was the first Russian quadrivalent inactivated subunit adjuvanted influenza and prevents against influenza A (H1N1 + H3N2) and influenza B (Yamagata and Victoria) for adults aged 18-60 and for children aged 6 or more (35, 36). According to the organization, the formulation has low reactogenicity and high efficacy. In fact, the use of a biodegradable and water-soluble adjuvant name Polyoxidonium(azoximer bromide) is responsible for cutting in three fold the antigen load compared to the traditional technology and it (the adjuvant) enhances the immune response (37). More recent work by Talayev and collaborators (2020) concluded that this quadrivalent formulation with Polyoxidonium induces the production of antibodies to all influenza hemagglutinins, enhances natural killer cells activity, induces Th2-polarized T-cell response and partial maturation of dendritic cells, which are stimulated to enhance inducible co-stimulator-ligant (ICOSL)’s expression. That is an important event to humoral immune response induction (38).

### 3.2 Patent Landscape - Human Vaccines

Our strategy search retrieved the global patent applications for vaccines against influenza A in humans for the 2010-2021 period resulted in 586 patent deposits, from a broad range of assignees/applicants in different countries as illustrated in Figure 4. The developed strategy also demonstrated that the leading priority country in number of applications was The United States (US) (with 197 documents), followed by China (153), Russia Federation (58) and South Korea (55). Based on the available data, the greatest number of documents were applied in 2010 (81 documents). In fact the average number of documents in each year was 49 applications.

**Figure 4.**
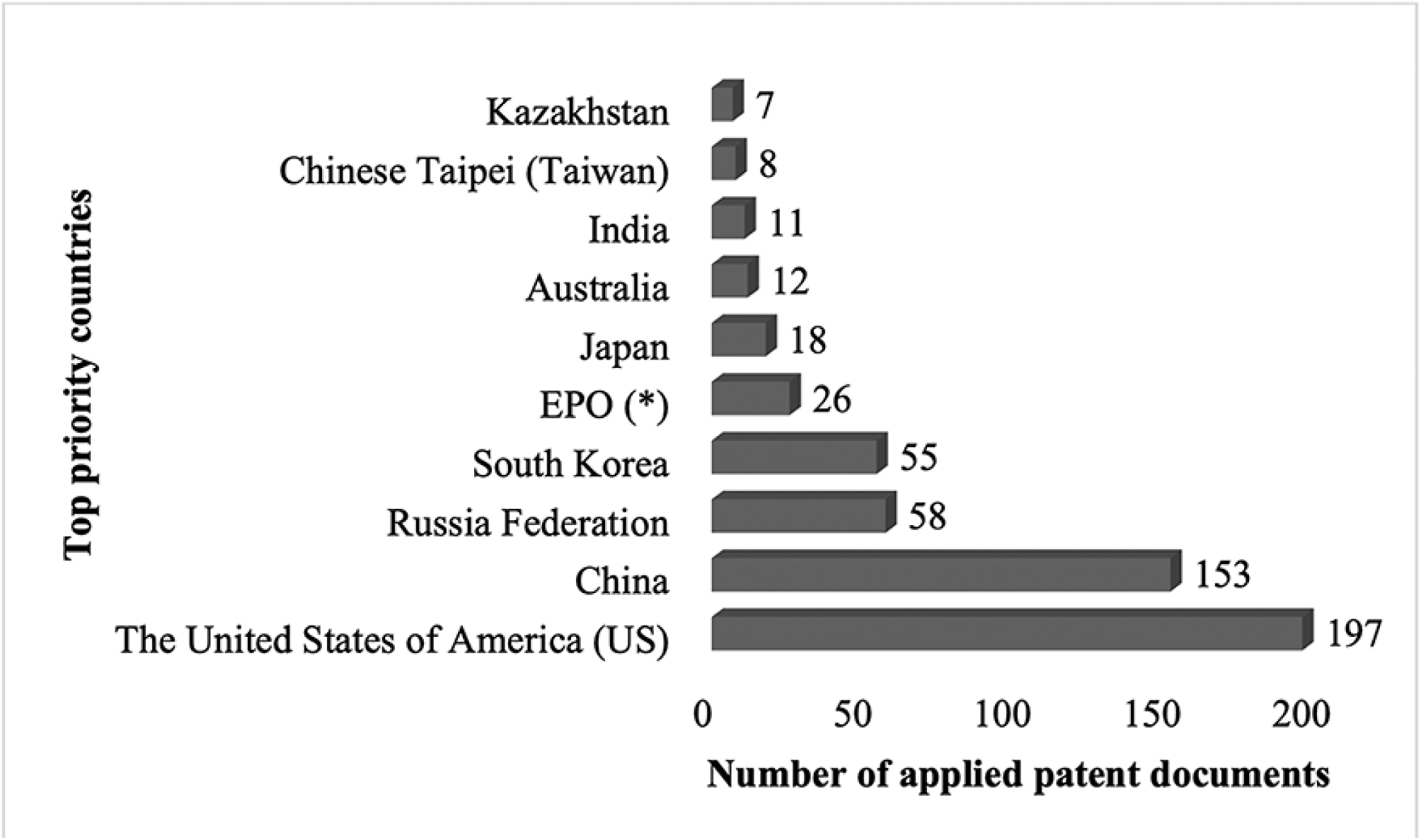
Top priority countries for vaccines against influenza A according to the number of applied patent documents during 2010-2021 period. (All the patents in different status were considered) Source: Elaborated by the authors from Derwent Innovation Index Database (22)

These results were obtained using the described methods to retrieve the applied documents from the leading applicants of vaccines against influenza A strains, i.e., H1N1, H2N2, H3N2 and H5N1. Based on the results (586 retrieved documents), the following observations are necessary: first, the leading applicant was from Russian Federation – THE INSTITUTE OF EXPERIMENTAL MEDICINE (with 29 applications), followed by the American Institution JANSSEN (11 documents) and the British Institution SEQIRUS UK LIMITED with 9 documents (Figure 5).

**Figure 5.**
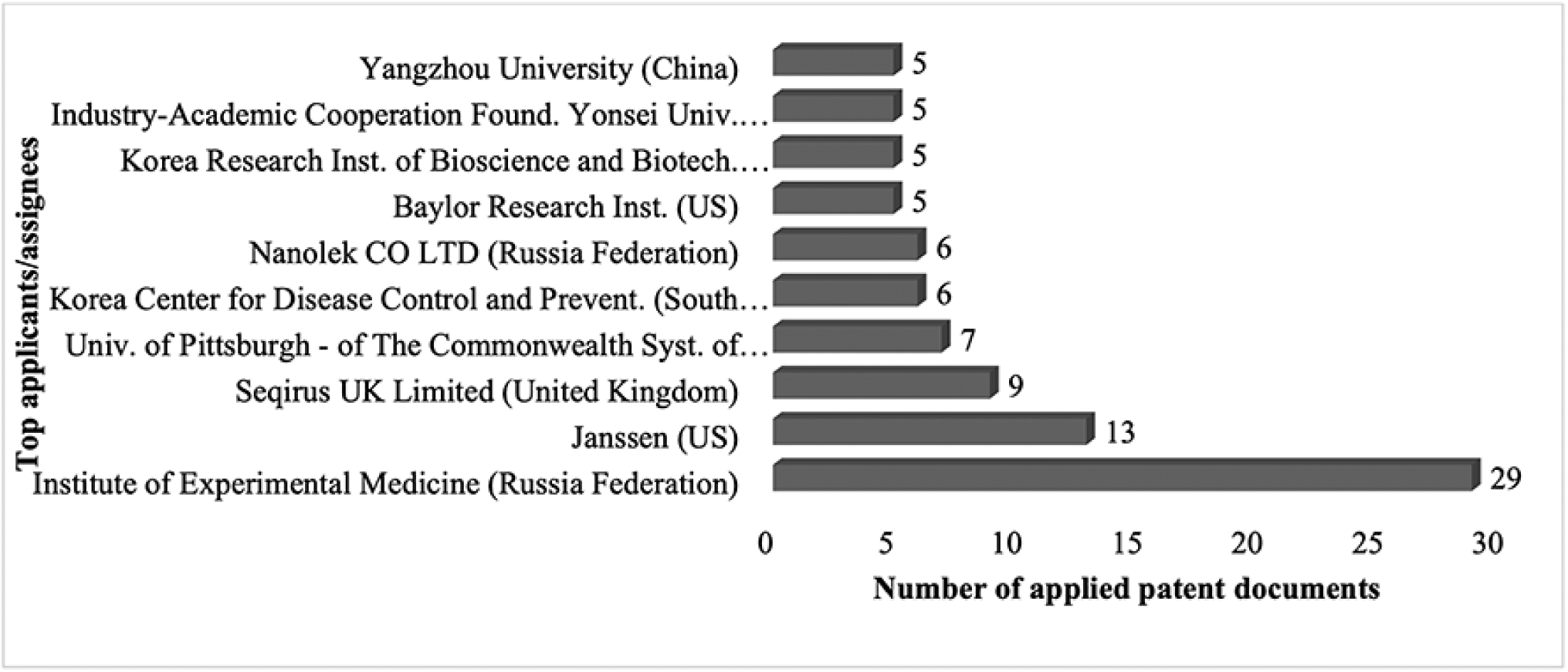
Top applicants/assignees for influenza A vaccine according to the number of applied patent documents during 2010-2021 period. (All the patents in different status were considered) Source: Elaborated by the authors from Derwent Innovation Index (22)

New Technologies for therapeutics and vaccines’ development to prevent and deal with future influenza pandemics are actively investigated not only to face and control next influenza pandemic strains, but also to assure the management of ongoing COVID-19 pandemic in humans (39). The method presented in this research was sufficiently general to describe how the patent documents were related to influenza A subtypes, i.e., H1N1, H2N2, H3N2, H5N1 and H7N9.In a second stage and after rigorous examination of these documents, we identified that most of them were related to vaccines to prevent/control/treat H1N1 influenza in humans. Moreover, we identified, as expected, a considerable number of documents related to more than one influenza A strain (Figure 6).

**Figure 6.**
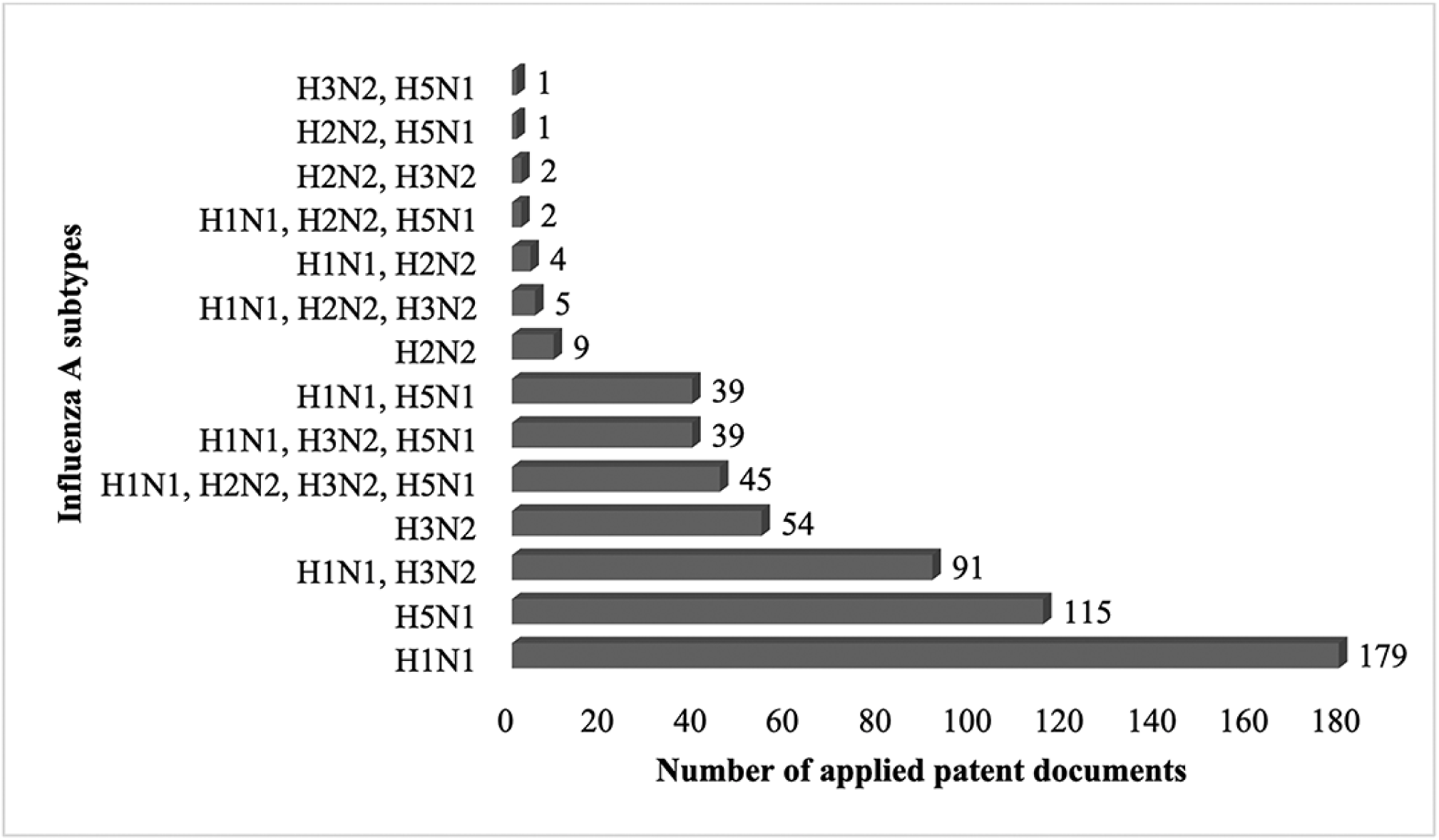
Influenza A subtypes cited in influenza vaccine documents Source: Elaborated by the authors from Derwent Innovation Index. (22) (*) For three documents, influenza strains were not described. The authors used the terms “influenza A” (2 documents) and “pandemic potential” (1 document) as it was described in the original applied patent documents. n: 586 (All the patents in different status were considered)

No further increase in the number of applications was observed for the other applicants/assignees. The results of our search have important global implications for Influenza pandemic preparedness and technological development of vaccines: the low number of documents for vaccines against influenza A strains by the leading applicants. This patent deposit scenario allows us to shed light on understanding the technological evolution of the three top companies in influenza vaccines development, i.e., INSTITUTE OF EXPERIMENTAL MEDICINE (from Russian Federation), PULIKE BIOLOGICAL ENGINEERING INC (from China) and JANSSEN (from US), indicating the strength of our methodological approach.

### 3.2.1 Institute of Experimental Medicine (Russia Federation)

The main achievements, including contributions from Russian the INSTITUTE OF EXPERIMENTAL MEDICINE may be summarized as follows. From the 29 retrieved applied documents, it was observed that most of the claimed vaccines were live intranasal, including live quadrivalent influenza ones from 2011 to 2019, i.e, almost all of the range study period (Figure 7).

**Figure 7.**
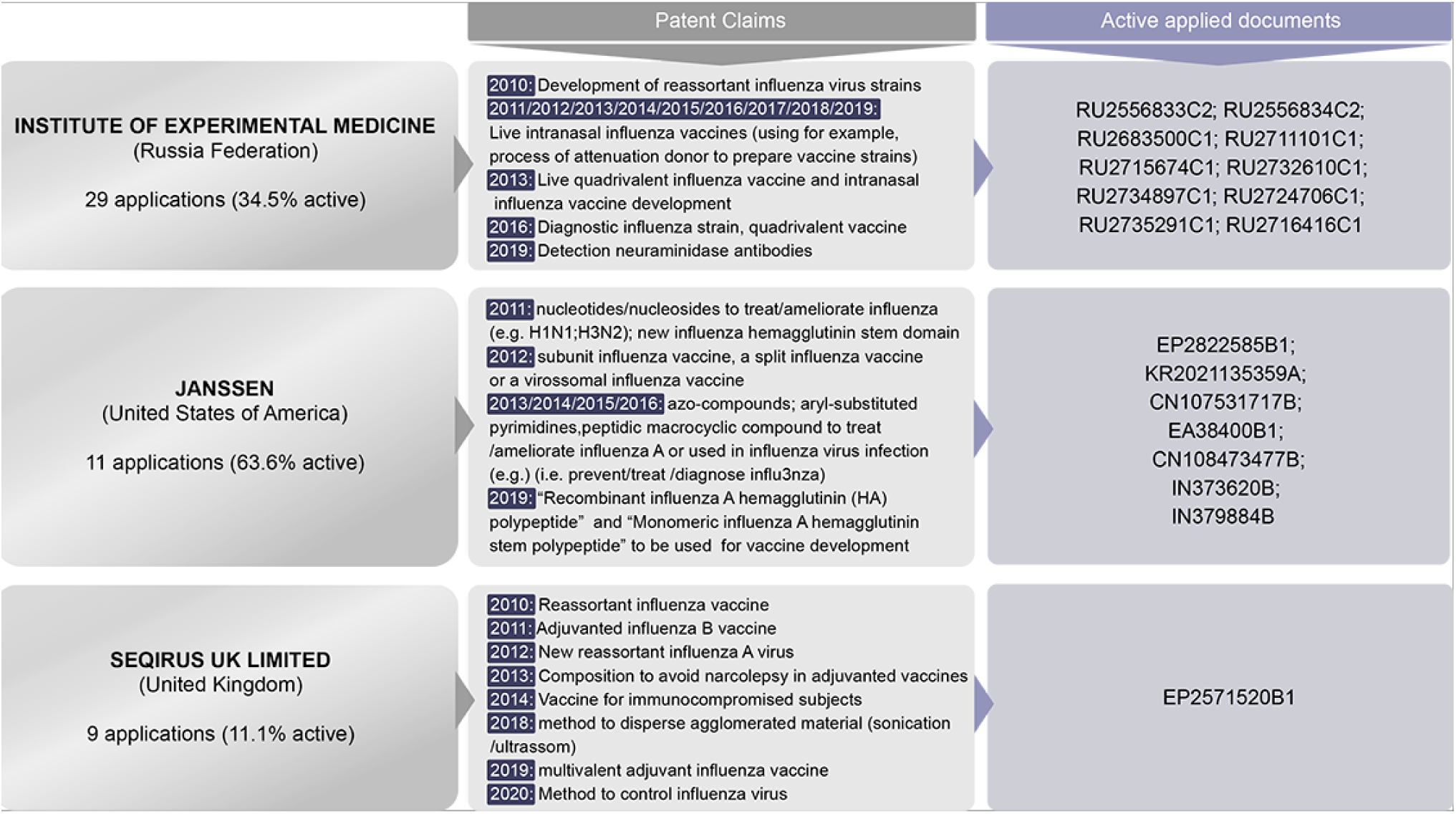
Main applicants’ chronological patent filings related to influenza A vaccines for human use Source: Elaborated by the authors from Derwent Innovation Index. (22, 40, 41, 42, 43, 44, 45, 46, 47, 48, 49, 50, 51, 52, 53, 54, 55, 56, 58) Note: All the patents in different status were considered (Supplementary Material I)

However, a closer examination of the results reveals different patent’s status that must be highlighted here. Once most of the applied Russian documents were inactive (17 documents – 58.6%) or discontinued (2 documents – 6.9%). So, the results here deviate significantly from the expected: a few numbers of active documents: 10 (only 34.5%) (Supplementary Material II). On the other hand the quality of the results is satisfactory, due to the documents offers information about the potential benefits of the use of biotechnology for the practical health service to prevent seasonal influenza. In 2011 the company developed from the Orthomyxoviridae family, it was obtained a new attenuated cold-adapted H1N1 strain and a reassortant H2N2 influenza (vaccine strain) to be used in an influenza intranasal vaccine for children and adults. The vaccinal strains (H2N2) are prepared by crossing epidemic viruses H1N1 (cold-adapted) and H2N2 strains. A key strength of the document lies within the fact that the attenuated H1N1 strain (cold-adapted) is new. This one is donor of attenuation to obtain vaccine strains. In fact, from this process it is possible to obtain strains of H2N2 (subtype A). Moreover, the H1N1 influenza strain has sensitivity to the preparations of adamantane row and temperature strip (PN: RU2556833C2) (40). In the same year (2011) another document is also related to a live intranasal influenza vaccine, where the vaccine strains were also obtained attenuation donor to prepare a H2N2 vaccine strain (PN: RU2556834C2) (41).

Once again, in 2018 the developed vaccine strain H3N2 was a candidate to produce a live intranasal influenza vaccine (PN: RU2683500C1) (42). In 2019, the close agreement of the results with the last applied documents suggested that the Russian Institute worked in the same line of research: the development of influenza vaccine strains, basically to live intranasal influenza vaccine for adults and children: H3N2 (PN: RU2711101C1 (43); RU2715674C1 (44) ; RU2732610C1 (45); RU2734897C1 (46)) or H1N1 (PN: RU2724706C1 (47); RU2735291C1 (48)). No applied documents were found in 2020 and 2021. However, it is clear the potential of this biotechnology to detect antibody to neuraminidase N2 in blood serum through a solid -phase inhibition reaction of neuraminidase activity using the reassortant strain H6N2 to detect neuraminidase antibodies (PN: RU2716416C1 (49)) (Figure 7).

### 3.2.2 Janssen (from US)

Based on the methodology used, once again, the number of documents retrieved from the second main applicant was low: 11 documents. From these, key findings emerged. In general and first, most of the documents claimed new compounds used to treat/ameliorate or to be used in influenza virus infection. To demonstrate this, examples of molecules can be presented: azo-compounds, aryl-substituted pyrimidines peptidic macrocycle compounds or nucleotides/nucleosides. Second, from the retrieved documents, most of them were active 7 (63.6%) were active, 1 application was discontinued (9.1%) and finally, 3 documents (27.3%) were discontinued (Supplementary Material I). From this scenario, we approach the low number of active documents with a broader perspective that the active applied documents are in fact the object of analysis to the present work, due to the interest in maintaining these documents by their applicants to protect the claimed products and methods. The company JANSSEN BIOPHARMA INC. (US) claimed in 2011 for a new substituted nucleotide and nucleoside compounds which are able inhibit viral replication and can be used treat/ameliorate paramyxovirus viral infection (e.g. pneumovirus infection) or orthomyxovirus viral infection (H1N1, H3N2, influenza A, B, C) or treat/ameliorate infections caused by parainfluenza, mumps, measles (e.g.) (PN: IN373620B (50)). JANSSEN VACCINES AND PREVENTION B.V., from Netherlands is part of JANSSEN PHARMACEUTICALS and has some applications in development of vaccines against influenza. The company claimed (priority year 2011) for a new influenza hemagglutinin stem domain used to treat influenza. The invention comprises hemagglutinin HA1/HA2 domains and “influenza hemagglutinin stem domain polypeptide is capable of broadly neutralizing antibody response and resistant to protease cleavage” (PN: IN379884B (51)).

The company JANSSEN VACCINES AND PREVENTION B.V. (Netherlands) claimed in 2012 (priority year) for a subunit influenza vaccine, split influenza vaccine or virosomal influenza vaccine which comprises the proteins neuraminidase (NA) and influenza hemagglutinin (HA) from three influenza strains (at least). In the interest of clarity, it may be mentioned that the vaccine is able to induce cross-protection against H5N1 strain (despite there was no NA/HA from H5N1 strain), due to the proteins from H3N2 and H1N1 and B strain of influenza (PN: EP2822585B1 (52)). In 2013 and 2015, the company claimed for aza-pyridone compounds to be used in pharmaceutical compositions to inhibit influenza virus’ replication such as H1N1, H3N2, H5N1, H7N9, Influenzas A and B (PN: KR2021135359A (53) and CN107531717B (54), respectively/JANSSEN BIOPHARMA INC). In 2014, the company claimed for a new influenza hemagglutinin stem with HA1 domain to induce immune response against current and future influenza virus strains. Moreover, it provides an effective and safe vaccine to stimulate robust neutralizing antibody response (PN: EA38400B1/JANSSEN VACCINES & PREVENTION B.V., Netherlands) (55). In 2016 the company claimed for aryl-substituted pyrimidines. The compounds are used in influenza virus infection. The compound was claimed to be used as a medicament (therapeutic agent: influenza vaccine, antiviral agent or both). The compound was tested against H1N1 through cell-based antiviral assay (PN: CN108473477B/JANSSEN SCIENCES IRELAND UC, Ireland (56)) (Figure 7).

### 3.2.3 Seqirus UK Limited (from United Kingdom)

SEQIRUS is a CLS company leader provider of pharmaceuticals and vaccines. Seqirus Pty Ltd is an affiliate from Australia and has applied some documents. It announced on March, 2022 the “Cell-based Flucelvax® Quad Influenza Vaccine”. It is approved for children two years age and older and it comprises a surface antigen, prepared in cell cultures and inactivated. In terms of quality of results, this approach delivers well. In fact, it is the only well-based/influenza vaccine available in Australia (57). As seen in Figure 7, the third main applicant (9 applications) claimed different products/methods, such as reassortant influenza vaccines/virus, multivalent adjuvanted vaccines or methods to disperse agglomerated material or to control the influenza virus (Supplementary Material). Our findings are somewhat surprising since, only one document from 2010 (PN: EP2571520B1 (58)) is active (11.1%), while 44.4% (4 documents) are discontinued and 44.4 (4 documents) are pending. Moreover, the active application consists in a contribution to existing knowledge and research is a set of techniques for vaccine production: The vaccine developed by SEQIRUS UK LIMITED contains per dose less than 10 ng residual host cell DNA. It is important to highlight that the production of reassortant virus was greatly increased due to (during virus production) the reduction of translation/transcription of the backbone strain’s neuraminidase/hemagglutinin genes. Furthermore, the claimed vaccine was a virosomal vaccine, surface antigen vaccine, a split virion vaccine or a whole virion vaccine. The viruseśstrains used were H1N1, H5N1, H5N3, H9N2, H2N2, H7N1 or H7N7.

## 3.3 Universal Vaccines

Results illustrated in Table 2 shows that universal vaccines are object of protection with individual advantages and technologies. In summary, the compositions cover a large subset of influenza virus strains. The adopted method attempted to retrieve 20 documents according to their status (Supplementary Material II). Table 2 summarizes the documents (in active status) about universal influenza vaccines used in human.

**Table 2.** Human universal influenza vaccines retrieved from the applied patent documents (from 2010-2021)

| HUMAN UNIVERSAL INFLUENZA VACCINES |  |  |  |  |  |
| --- | --- | --- | --- | --- | --- |
| Patent document number (earliest priority year) | Applicant/ assignee (Country) | Vaccine/ Antibodies | Influenza Strains | Considerations | Advantages (e.g.) |
| KR1382244B1 (2012) (71) | MOGAM BIOTECHNOLOGY RESEARCH INSTITUTE (Republic of Korea) | T cell-based universal vaccine | *“broad-spectrum defense against various hetero-subtypic variants of influenza viruses”* | The universal flu vaccine contains nucleoprotein gene/polymerase acidic gene | - The vaccine expresses a broad-spectrum against influenza variants (many hetero-subtypic variants). |
| CN103505725B (2013) (72) | CSG MICROBIAL IMMUNE PREPARATION ENGINEERING CENTER NANTONG CO. LTD (China) | New cross-protective effect influenza vaccine | *“H1N1 influenza virus, human influenza virus, swine influenza virus or avian influenza virus”* | The vaccine contains *“a common heat shock glyco protein 96”* | -The universal influenza vaccine exhibits cross protection performance. |
| CN103948942B (2014) (73) | QINGDAO AGRICULTURAL UNIVERSITY (China) | Influenza virus universal gene vaccine | Prevention against *“the occurrence of various influenza virus diseases”* | The vaccine *“is formed by cloning hemagglutinin 2 gene of H1N1 virus strain of influenza with pCAGGS eukaryotic expression vector”* | - The vaccine induces fine cell immune and humoral responses.<br>-The vaccine exhibits less adverse reactions.<br>- The vaccine is able to prevent the disease against various influenza disease |
| US10456464B2 (2014) (74) | NITTO DENKO CORPORATION (Japan) | The vaccine the antigen and antithrombotic drug to induce humoral immunity | H1N1, H3N2, Influenza B virus, Influenza B virus and other antigens, such as *“live attenuated mumps virus, live attenuated measles virus, live attenuated rubella virus, tetanus toxoid-conjugated haemophilus influenzae type b polysaccharide”*, etc | The vaccine can be used universally due to the induction of humoral immunity against many antigens | The composition is not invasive for the patients |
| RU2618918C2 (2015) (75) | GAMALEYA EPIDEMIOLOGY MICROBIOLOGY FED | Universal anti-infectious vaccine | H1N1, H2N3, H3N2, H5N2, H9N2 | The vaccine is comprised by recombinant pseudo-adenovirus particles | The composition has a broad-spectrum of antiviral activity |

HUMAN UNIVERSAL INFLUENZA VACCINES
| Patent document number (earliest priority year) | Applicant/ assignee (Country) | Vaccine/ Antibodies | Influenza Strains | Considerations | Advantages (e.g.) |
| --- | --- | --- | --- | --- | --- |
| US10953086B2 (2018) (76) | INDUSTRY-ACADEMIC COOPERATION FOUNDATION YONSEI UNIVERSITY (Republic of Korea) | Universal Influenza vaccine | It produces immune response “against at least one HA protein (HA group 1) such as H1, H2, H5, H6, H8, H9, H11, H12, H13, and H16; and at least one HA protein (HA group 2) selected from the group consisting of H3, H4, H7, H10, H14, and H15” and against “heterologous influenza viruses” | The vaccine uses cold-adapted live-attenuated virus | The vaccine ensure protection and can exhibit cross-protective effect against influenza virus strains |
| KR2211378B1 (2019) (77) | KOREA RESEARCH INSTITUTE OF BIOSCIENCE AND BIOTECHNOLOGY (Republic of Korea) | Influenza vaccine | H1N1, H1N2, H2N2, H3N1, H3N2, H5N1 and H5N8 | The vaccine induces immune response against new mutant influenza virus and influenza virus subtypes | The fusion protein can produce neutralizing antibodies against influenza subtypes and swine mutant influenza virus |

Source: Elaborated by the author from Derwent Innovation Index (22)
Note: Patent status – active

## 4 Discussion

The results of our search have important global implications for Influenza pandemic preparedness and the technological development of influenza vaccines: the low number of documents for innovative vaccines against potentially-pandemic influenza A strains by the leading applicants. From 49 deposits in the search period (2010–2021), only 18 products in active status (37%) were retrieved. In addition, most of them referred only to incremental technologies. This scenario is concerning and alerts to the urgent need to prevent this disease worldwide, considering its pandemic potential and high mortality associated with influenza A, mainly in the elderly and children.

### 4.1 Vaccine scale-up: bottlenecks

These results indicate that it will be very difficult to scale-up these vaccines in the case the world needs them. Current egg-based processes for vaccine production are too laborious and difficult to scale-up in the case of a pandemic. Concerning the use of mRNA technology, there are still many open questions on the duration and level of protection mRNA vaccines might induce. Vaccinating just birds, on the other hand, is not easy nor sufficient: if not carried out properly, the virus could continue to circulate at a low level, actually increasing the chance of mutations and spread. Universal Influenza vaccines will be thus key for HPAI pandemic preparedness. However, our results identified few documents for these vaccines (20 in the search period) and indicate that they are still in early stages of development, facing many gaps and challenges to be introduced into the market. In addition, it is very difficult to predict the impact immunological memory would have on the development of a broad-spectrum vaccine due to original antigenic sin mechanism.

### 4.2. Applicantś strategies

Considering the main applicants for human flu vaccines: the Russian Institute (Institute of Experimental Medicine) focused its research (among other strategies) in the development of reassortant influenza virus strains and quadrivalent vaccines and vaccines for intranasal use, as observed in the following applied documents: PN: RU2556834C2 (59), RU2683500C1 (42), RU2711101C1 (43); RU2715674C1 (44); RU2732610C1 (45); RU2734897C1 (46) and RU2735291C1 (48).

The American company (Janssen) with a more diversified portfolio acted in different strategies in the development of vaccines by use of new compounds (azo-compounds – PN: KR2021135359A (53) and PN: CN107531717B (54) and aryl-substituted pyrimidines – PN: CN108473477B (56).

Concerning the documents applied by the Institute of Experimental Medicine, the documents which claimed for intranasal vaccine (PN: RU2556834C2 (59)) was selected. From the American company Janssen, these innovations were observed in a new influenza hemagglutinin stain domain polypeptides (PN: IN379884B (51)) which was also a retrieved innovation from Janssen.

This patent deposit scenario allows us to shed light on the urgent need to accelerate the global effort towards more effective and long-duration influenza vaccines, supporting vaccine innovation and the scientific and technological development of a universal pan-influenza vaccine.

### 4.3 Technological issues: mRNA and universal vaccines

Our results also indicate that the number of patent documents for highly pathogenic Influenza A human vaccines, besides being still very low, are mostly referred to incremental innovations, in spite of significant recent breakthroughs in cell-based and recombinant vaccines. In fact, an effective influenza pre-pandemic vaccine is difficult to obtain, due to the impossibility to develop the vaccine against the correct virus subtype to the next pandemic. New technologies are available to provide protection against influenza strains, such as “nucleoside-modified messenger RNA (mRNA)–lipid nanoparticle vaccine” which is able to act in a multivalent way, i.e., the formulation can encode hemagglutinin antigens for influenza A subtypes (20 known) and B lineages (60).

Nevertheless, in spite of recent studies using mRNA technology for these new influenza vaccines, they have not advanced much yet. The main issue constraining mRNA vaccine development for pathogens that mutate fast, such as the potentially pandemic Influenza H7N9 and H5N1 strains, is producing the correct antigen. The problem with Influenza vaccine and other vaccines for potentially pandemic or pandemic diseases such as HIV is that so far researchers have not been able to engineer the antigen to present the correct pan-neutralizing epitopes.

Consequent to these innovation constraints, the current Influenza vaccines examined in our study have important limitations: in addition to their low efficacy and effectiveness, due to the high mutation and viral antigenic variation, the protection conferred is of short duration. It is also important to underline the need for new adjuvant technologies for longer Influenza vaccine protection and engineering the antigens to expose to the immune system the pan-neutralizing epitopes in their correct conformation. Finally, there is also a need to accelerate the development of more Influenza vaccines with nasal application, besides Flumist*. Finally, a novel strategy of combined Influenza-COVID-19 vaccines should be developed, and some efforts have already started in this direction.

Considering this scenario, the development of innovative vaccine technologies for large scale production is urgent for improving vaccine efficacy for seasonal influenza and for global vaccine pandemic preparedness. The complexity of influenza viral evolution, requiring annual shots due to emergence of new variants evidences the need for urgently accelerating funding and incentives towards innovative vaccines and a universal pan-influenza vaccine against all Influenza strains. A universal vaccine would be very important if it can combine protection for different antigens and viral mutations and confer a long protection, which seems still very difficult. The COVID-19 pandemic has highlighted these limitations and provides a critical drive for vaccine development and regulatory innovation.

### 4.4 Human and veterinary vaccines: One Health

Accelerating vaccine development for HPAI, reviewing current time frames, has proved thus to be a critical issue. The development process for a human vaccine can take between 10 and 20 years. By contrast, veterinary vaccines usually take between 3 and 6 years, according to the country where the product is registered (61). Veterinary vaccines must also be urgently improved and object of accelerated Research, Development and Innovation (RDI). Numbers from the World Organization for Animal Health (WOAH) indicate that the huge impacts of HPAI and other zoonotic diseases on wildlife could potentially lead to a devastating effect on the biodiversity of global ecosystems. This scenario alerts to the need for effective and immediate response efforts to zoonotic diseases and for coordinated multi-sectorial prevention as important tools to the control of HPAI and reinforces the One Health concept: 1. 80% of pathogens of bioterrorism concern are originated in animals; 2. 75% of the emerging pathogens in human are from animals and finally, 3. 60% of pathogens that can cause diseases in human are from wild or domestic animals. The world faces COVID-19 pandemic, which resulted from a virus of potential animal origin (62), with 762.791.152 confirmed cases and 6.897.025 deaths globally. Fortunately, a total of 13.340.275.493 COVID-19 vaccine doses have been administered (63), the best and current example of innovation, technological development for pandemic preparedness for future generations. Vaccine Preparedness for Influenza pandemic should be thus certainly inspired in these COVID-19 pandemic lessons and success story.

### 4.5 Vaccine Preparedness model

Differently from the rapid global vaccine response to COVID-19 pandemic, in less than six months, due to previous developments and strong funding mechanisms from the US government (Warp Speed Operation) and international agencies, our results indicate that in the case of a future HPAI outbreak or pandemic, this global vaccine response might probably be much slower and weaker. Even if funding mechanisms similar to COVID-19 response are immediately put in place for HPAI vaccine development, there are still many technological and production platform gaps and bottlenecks to be overcome. These constraints in vaccine development contrast with reports of new HPAI human cases, now also in the American region. On March 2023, the second H5N1 human case in South America has been reported by the government of Chile. The first case in this region had been reported in Ecuador in December 2022 in the US, one case has been already reported in April 2022 and was the first case in the country (64). Recently, on March 27 2023 the National Health Commission of the Peoplés Republic of China notified the death from Avian Influenza (H3N8) of a 56-year-old female from Guangdong province hospitalized for severe pneumonia on March 3 and subsequently died on March 16 2023. No close contact of the case developed an infection or symptoms or illness at the time of reporting. The results of testings in environmental areas from the residence and the wet market where the patient spent time before the onset of illness showed samples positive for influenza A in the wet market (H3) (65).

This concerning scenario highlights the urgent need for a Vaccine Preparedness model, supported by extensive Genomic, Antigenic and Epidemiological Surveillance, an Innovation Fund and Public-Private Partnerships, encompassing an effective global response. This model should be based on more sensitive surveillance, more relevant pre-clinical models and a prototype vaccine bank ready for production and clinical trials. Incentives to accelerate innovation and technology transfer for multipatented vaccines, such as “patent pools” should also be provided (66). We strongly support WHO in coordinating a Global Initiative for Pandemic Preparedness, in partnership with the Coalition for Epidemic Preparedness Innovations (CEPI) and other partners: governments, NGOs and stakeholders. Some initiatives, such as the Influenza Vaccines Research and Development (R&D) Roadmap (IVR) were created with this perspective, through an extensive international stakeholder engagement process, to promote influenza vaccine R&D (67) (Figure 8).

**Figure 8.**
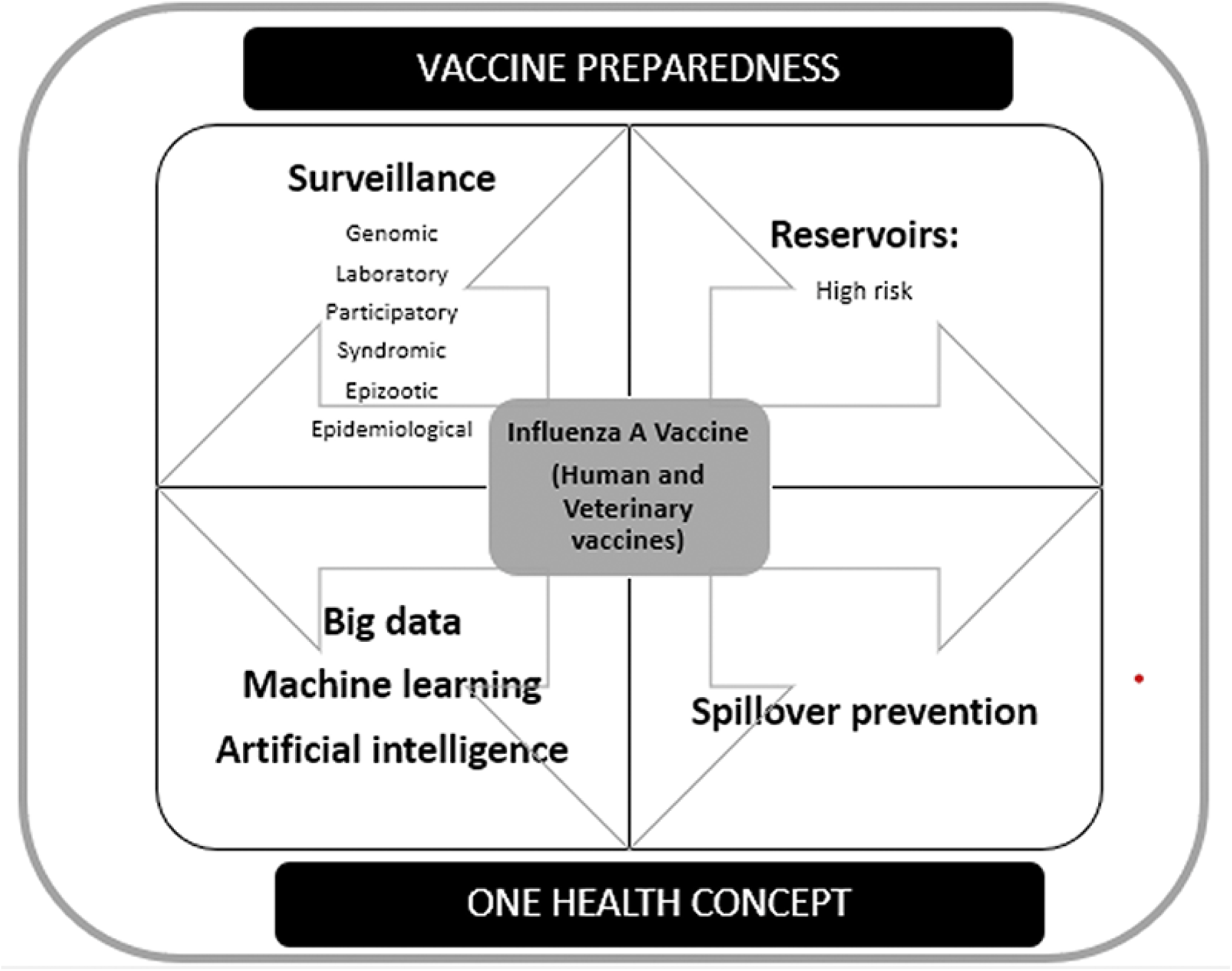
Towards a new global eco-social influenza pandemic preparedness system. Source: elaborated by the authors.

In conclusion, our results provide evidence on the main gaps and bottlenecks for HPAI vaccine development and scale-up in the case of a pandemic, stressing the urgent need to speed-up vaccine development in a global Vaccine Preparedness model, based on a comprehensive One Health conceptual framework. This model should integrate a Global Strategic Plan to Accelerate Innovation, Technological Development and Production of Influenza Vaccines. These strategies will require clear priorities, innovative funding and new business models, integrating academia, pharmaceutical enterprises and governments.

## 6 Author Contributions

CP, EM, AA, AO and AH contributed to conception and design of the study. AA, AO, SS and FM organized the database and performed the statistical analysis. CP, AA and AO wrote the first draft of the manuscript. AA, EM and MS wrote sections of the manuscript. All authors contributed to manuscript revision, read, and approved the submitted version.

## Data Availability

All data produced in the present work are contained in the manuscript

## 7 Acknowledgments

The authors thank Bio-Manguinhos of Oswaldo Cruz Foundation, the School of Chemistry of the Federal University of Rio de Janeiro and the National Institute of Industrial Property for the support. The authors gratefully acknowledge the financial support of University of Pittsburgh. We also thank Rafael Cavalcante dos Santos, M. Sc., School of Chemistry, Federal University of Rio de Janeiro for the figures and final art.

## Footnotes

1 Dr Peter Palese – Department of Microbiology at Icahn School of Medicine (New York City) (15).

2 CCDI is a Clarivate Database focused on drug and pharma development where it is possible to obtain integrated pharmacological, chemical and biological data of the formulation of interest. The search was carried out on September 6, 2022 (17).

3 In Drugs & biologics (Advanced search): Condition – Influenza A and Product Category – vaccines. First filter: Year launched/registered: from 2010 to 2021; second filter: condition: influenza A and third condition: development status: launched.

4 Derwent World Patent Index (from Clarivate) Manual is a hierarchical indexing system to be used as an analysis tool and a patent retrieval in patent searching. The strains H1N1, H2N2, H3N2 and H5N1 were chosen according to their pandemic potential (20).

5 Active: “Granted patent in force. If an application has been Granted, all published documents associated with the application have a status Active”. Discontinued: “Application discontinued, withdrawn or rejected, i.e. discontinuation before Grant”. Pending: “Application is pending (Pending: IP Right has not been granted yet and application is in either filing, examination, pre-grant stage)”. Inactive: “Granted patent not in force because of lapse, non-fee payment, etc. The patent hasn’t reached the term date and can be revived” (The Lens (18), Indian Patent Advanced Search System (19)).

6 Derwent Innovations Index (from Clarivate) covers over 14.3 million basic inventions. The documents are from almost 60 authorities in worldwide. The search was carried out on 2 February 2022, and it will retrieve dead/alive applied patent documents (22).

7 The terms were chosen from the following references: World Health Organization (23), Medical Subjects Headings (Mesh) from National Library of Medicine (24) and Darricarrère N et al. (25)

8 Here, influenza B vaccine were also retrieved (e.g. quadrivalent vaccines)

9 Launched: “The drug is being marketed” (26).

