## Supplemental Tables 1 and 2 for "Highly Pathogenic Avian Influenza Vaccines: challenges for innovation, technological development and pandemic preparedness"

### *Supplementary Material*

#### 1 Supplementary Tables

##### 1.1 Supplementary Table 1. Human vaccines: applied documents and their patent status from the main applicants

| Applicants (Country/ total of retrieved documents) | **Patents' status** | | | |
|---|---|---|---|---|
| | According to The Lens / Indian Advanced Search System (*) | | | |
| | Active | Discontinued | Pending | Inactive |
| | n (%) | n (%) | n (%) | n (%) |
| THE INSTITUTE OF EXPERIMENTAL MEDICINE (Russia/29) | RU2556833C2 (1)<br>RU2556834C2 (2)<br>RU2683500C1 (3)<br>RU2711101C1 (4)<br>RU2715674C1 (5)<br>RU2732610C1 (6)<br>RU2734897C1 (7)<br>RU2724706C1 (8)<br>RU2735291C1 (9)<br>RU2716416C1 (10) | RU2013149103A (11)<br>RU2016146943A (12) | | RU2416640C1 (13)<br>RU2416641C1 (14)<br>RU2422517C1 (15)<br>RU2422518C1 (16)<br>RU2422519C1 (17)<br>RU2464310C1 (18)<br>RU2464311C1 (19)<br>RU2532844C2 (20)<br>RU2563352C2 (21)<br>RU2587629C1 (22)<br>RU2013149106A (23)<br>RU2606026C1 (24)<br>RU2605314C1 (25)<br>RU2606019C1 (26)<br>RU2627188C1 (27)<br>RU2625024C1 (28)<br>RU2653388C1 (29) |

| | | | | |
|---|---|---|---|---|
| | **10 (34.5)** | **2 (6.9)** | **0 (0.0)** | **17 (58.6)** |
| JANSSEN (USA/11) | EP2822585B1 (30)<br>KR2021135359A (31)<br>CN107531717B (32)<br>EA38400B1 (33)<br>CN108473477B (34)<br>IN373620B (35) (*)<br>IN379884B (36) (*) | IN201727012541 (37) (*) | US20200048308A1 (38)<br>CN113597428A (39)<br>WO2021043869A1 (40) | |
| | **7 (63.6)** | **1 (9.1)** | **3 (27.3)** | **0 (0.0)** |
| SEQIRUS UK LIMITED (United Kingdom/9) | EP2571520B1 (41) | US20190247489A1 (42)<br>US20200000909A1 (43)<br>US20190184007A1 (44)<br>AU2021202331A1 (45) | JP2021531339A (46)<br>WO2022005893A1 (47)<br>AU2020228151A1 (48)<br>AU2019302562A1 (49) | |
| | **1 (11.1)** | **4 (44.4)** | **4 (44.4)** | **0 (0.0)** |

Legends:

***Active**: “Granted patent in force. If an application has been Granted, all published documents associated with the application have a status Active”*

***Discontinued**: “Application discontinued, withdrawn or rejected, i.e. discontinuation before Grant”*

***Pending**: “Application is pending (Pending: IP Right has not been granted yet and application is in either filing, examination, pre-grant stage)”*

***Inactive**: “Granted patent not in force because of lapse, non-fee payment, etc. The patent hasn't reached the term date and can be revived”.*

Source: Elaborated by the authors from The Lens and Indian Patent Advanced Search System. The search was carried out on November 21st, 2022. (50, 51)

**Supplementary Table 2. Universal Vaccines: applied documents and their patent status from the main applicants**

| Applicants (Country/ total of retrieved documents) | **Patents' status** | | | |
|---|---|---|---|---|
| | According to The Lens / Indian Patent Advanced Search System (*) | | | |
| | Active | Discontinued | Pending | Inactive |
| | n (%) | n (%) | n (%) | n (%) |
| Ertl Hildegund C. J., and Zhou Dongming (US) | | US 20130209512 (52) A1 | - | |
| Temasek Life Sciences Laboratory Ltd (Singapore) | | IN201203735P2 (53) (*) | | |
| The Secretary of State for Health (United Kingdom) | | IN201401597P1 (54) (*) | | |
| Streck INC. (US) | | US20180243405A1 (55) | | |
| The Wistar Institute of Anatomy and Biology (US) | | US20140377295A1 (56) | | |
| Mogam Biotechnology Research Institute (Republic of Korea) | KR1382244B1 (57) | | | |
| Henan Agricultural University (China) | | | | CN102600466B (58) |
| CSG Microbial Immune Preparation Engineering Center Nantong CO. LTD (China) | CN103505725B (59) | | | |
| Sanofi Pasteur INC (US) | | US20170121373A1 (60) | | |
| Qingdao Agricultural University (China) | CN103948942B (61) | | | |
| Nitto Denko Corporation (Japan) | US10456464B2 (62) | | | |
| Gamaleya Epidemiology Microbiology FED | RU2618918C2 (63) | | | |
| Sun Yat-Sen University (China) | | CN105950646A (64) | | |
| The Board of Regents of The University of Texas System (US) | | | US20190351046A1 (65) | |
| Industry-Academic Cooperation Foundation Yonsei | US10953086B2 | | | |

| | | | | |
|---|---|---|---|---|
| University (Republic of Korea) | (66) | | | |
| Xu Jian Qing (China), Zhang Xiao Yan (China), Wang Jing (China) Zhu Ling Yan (China),Lubit Beverly W (US) | | | WO2021141758A1 (67) | |
| Korea Research Institute of Bioscience and Biotechnology (Republic of Korea) | KR2211378B1 (68) | | | |
| University of Manitoba (Canada) | | | WO2021077215A1 (69) | |
| The Trustees of The University of Pennsylvania; Icahn School of Medicine at Mount Sinai (US) | | | WO2021202734A3 (70) | |
| Massachusetts Institute of Technology (US) | | US20210379179A1 (71) | | |
| **Total 20 (100%)** | **7 (35.0)** | **8 (40.0)** | **4 (20.0)** | **1 (5.0)** |
